# Trajectories of blood-based protein biomarkers in chronic traumatic brain injury

**DOI:** 10.1101/2025.02.16.25322303

**Authors:** Amelia J. Hicks, Jay Plourde, Enna Selmanovic, Nicola L. de Souza, Kaj Blennow, Henrik Zetterberg, Kristen Dams-O’Connor

## Abstract

Blood-based protein biomarkers may provide important insights into the long-term neuropathology of traumatic brain injury (TBI). This is urgently required to identify mechanistic processes underlying post-traumatic neurodegeneration (PTND); a progressive post-recovery clinical decline experienced by a portion of TBI survivors. The aim of this study was to examine change over time in protein levels in a chronic TBI cohort. We selected six markers (Aβ_42_/Aβ_40_, GFAP, NfL, BD-tau, p-tau231, and p-tau181) with known importance in acute TBI and/or other neurodegenerative conditions.

We used a longitudinal design with two time points approximately 3.5 years apart on average (SD 1.34). Proteins were measured in plasma using the ultrasensitive Single molecule array technology for 63 participants with mild to severe chronic TBI (sustained ≥ 1 year ago; M 28 years; SD 16.3 since their first blow to the head) from the Late Effects of TBI study (48% female; current age M 52 years; SD 13.4). Multivariate linear mixed effect models with adjustments for multiple comparisons were performed to examine trajectories in proteins over time with age and age squared as covariates. A series of sensitivity analyses were conducted to account for outliers and to explore effects of key covariates: sex, *APOE* ε4 carrier status, medical comorbidities, age at first blow to the head, time since first blow to the head, and injury severity.

Over an average of 3.5 years, there were significant reductions in plasma Aβ_42_/Aβ_40_ (β = −0.004, SE = 0.001, t = −3.75, q = .001) and significant increases in plasma GFAP (β = 12.96, SE = 4.41, t = 2.94, q = .01). There were no significant changes in NFL, BD-tau, p-tau231, or p-tau181.

Both plasma Aβ_42_/Aβ_40_ and GFAP have been associated with brain amyloidosis, suggesting a role for Aβ mis-metabolism and aggregation in the long-term neuropathological consequences of TBI. These findings are hypothesis generating for future studies exploring the diverse biological mechanisms of PTND.

---

Traumatic brain injury (TBI) is now recognized as a chronic and dynamic health condition.^1,2^ Following a period of recovery, many individuals^3^ experience ongoing but stable functional disability,^4–7^ cognitive impairment,^5,8–13^ poor emotional health,^3,5,7,10,14–17^ and neurobehavioral dysregulation^3,18^ with lifelong implications on quality of life. Approximately one third of individuals, however, experience post-recovery decline that is not accounted for by age.^19–21^ The term ‘post-traumatic neurodegeneration’ (PTND) has been employed to distinguish chronic but stable post-TBI impairments from the progressive decline experienced by some.^22–27^ The notion that a subset of TBI survivors may experience a decline in clinical function years after injury has been recognized for decades through work highlighting the elevated rates of dementia following TBI.^28–30^ Efforts to classify post-TBI dementia within existing neurodegenerative diseases, mainly Alzheimer’s disease (AD),^31–58^ have yielded mixed and inconclusive results (Parkinson’s disease,^32,34,59–61^ Frontotemporal dementia,^32,62^ Amyotrophic Lateral Scleorsis,^63,64^ and Vascular Dementia).^65^ PTND is now considered a multiple etiology dementia (MED) in the AD and Alzheimer’s disease-related dementia (AD/ADRD) framework,^66^ that appears to have distinct clinical presentations.^40,65,67–69^ The pathologies driving PTND remain unknown,^25,26^ and the long-term neuropathological consequences of TBI more broadly have been understudied. Knowledge advancement has been impeded by a dearth of post-mortem tissue from long-term survivors of TBI, which was historically been available for individuals who had died acutely after injury.^70–82^

In recent years, there has been significant investment in blood biomarkers research,^83,84^ which unlike PET imaging with radiotracers and cerebrospinal fluid analyses requiring lumbar puncture, presents a cost-effective and minimally invasive method to capture pathology during life. To date, TBI blood biomarkers have almost exclusively focused on acute TBI diagnosis and prognostication,^85–87^ with several promising markers identified: S100 calcium binding protein B (S100B), ubiquitin carboxy-terminal hydrolase L1 (UCH-L1), glial fibrillary acid protein (GFAP), and neurofilament light (NfL).^86,88^ A recent systematic review found only 30 studies investigating blood biomarkers in the chronic stages (≥1 year post-injury).^89^ The most robust evidence from studies with TBI sample sizes ranging from n=22-298, identified elevations in select proteins compared with controls, even in those with mild TBI.^90,91^ This includes inflammatory markers (12 months: eotaxin-1, IFN-y, IL-8, IL-9, IL-17A, MCP-1, *MIP-1*β, FGF-basic, and TNF-α^91^; 9 years: IL-6^90^), GFAP (12 months^92–94^ and M 7 years^95^), UCHL-1 (M 22 years^58^), p-tau181 (M 22 years^58^), and neurodegenerative-disease associated protein variants of amyloid-β (Aβ), TAR DNA-binding protein 43 (TDP-43), α-synuclein, and tau (M 29 years^57^).

Notably, most TBI blood biomarker studies have been cross-sectional^96^ precluding characterization of trajectories over time. This is most notable for chronic TBI cohorts, for which there is limited examination of trajectories beyond 12 months. Within the first 12 months post-injury, most evidence suggests trajectories of decrease over time for brain derived tau (BD-tau; 0 days – 12months;^97^ 7 days – 12 months^97^), p-tau231 (0 days – 12months;^97^ 7 days – 12 months^97^), NfL (7 days – 12 months;^97^ 3 – 12 months;^98^), and total tau (0 days – 12months;^97^ 7 days – 12 months;^97^ 3 – 12 months^98^). Important questions remain about whether changes in these blood biomarkers can be detected years after injury and their potential role in long-term clinical dysfunction. Evidence of ongoing changes in protein levels in the chronic TBI period would be extremely informative for elucidating pathological processes contributing to PTND.

To draw accurate conclusions about biomarker trajectories in chronic TBI, it is essential that we account for known associations with key demographic variables. Failure to consider contributions from these variables may lead to incorrect interpretations about the role of TBI in protein change over time. Increasing age has been consistently associated with several commonly examined blood-based protein biomarkers (e.g. Aβ42,^99^ Aβ40,^99^ Aβ42/40,^99^ NfL,^99,100^ T-tau,^99,101^ p-tau,^102^ inflammatory markers^103^). Sex may also impact levels of some proteins, with supporting evidence in non-TBI^104^ and TBI cohorts^91,98,105–108^. Likewise, *APOE* ε4 genotype has been robustly associated with levels of several proteins measured in blood, Aβ,^109^ GFAP,^110^ NfL,^111^ and the progression of pathological burden over time.^112^ Medical health conditions may also impact some protein levels. This is particularly important in TBI cohorts, in which comorbidity of multiple medical conditions is common. ^113–115^ Several conditions that are commonly comorbid with TBI^116,117^ have been independently associated with increased levels of select blood-based proteins including diabetes,^118^ cardiovascular conditions,^118^ and cerebrovascular conditions.^119^

The aim of the current study was to examine change over time in protein levels in a longitudinal cohort of individuals living with TBI. We selected six markers with known importance in acute TBI and/or other neurodegenerative conditions to test the hypotheses that: (1) there will be a reduction in plasma Aβ_42_/Aβ_40_ levels over time, and (2) an increase in plasma levels of GFAP, NfL, BD-tau, p-tau231, and p-tau181.

## Methods

### Participants

Participants were enrolled in the Late Effects of Traumatic Brain Injury Study (LETBI).^120^ The LETBI Study is an ongoing multi-center prospective longitudinal brain donor program. The aims of the LETBI study are to identify the clinical characteristics, in-vivo biomarkers, and postmortem neuropathology of PTND. Participants complete longitudinal study visits for clinical assessment, biosample collection, and neuroimaging, and make known their wishes for brain donation. Study visits are scheduled approximately every 2-3 years. LETBI participants are recruited from TBI research registries at the Icahn School of Medicine at Mount Sinai (ISMMS), The University of Washington, Massachusetts General Hospital, Indiana University, Kessler Institute for Rehabilitation, and New York University, as well as from community-based settings. This study reports on data collected from the ISMMS site only.

Eligible participants were: (1) aged 18 years or older, (2) has sustained at least two mild TBIs, or at least one complicated mild, moderate, or severe TBI (defined as a blow to the head resulting in a period of altered mental status or unconsciousness)^121^ (3) at least 1 year post first head trauma exposure, and (4) English speaking. Participants are not removed from the LETBI study if they sustain a subsequent head trauma during study participation, as recurrent head trauma exposure is common^122–124^ and has important implications for long-term outcomes.^125,126^

All participants (or their proxies) provided written informed consent before any study procedures as per protocols approved by the ISMMS program for the protection of human subjects. *Head trauma exposure ascertainment*

The Brain Injury Screening Questionnaire (BISQ)^127,128^ was used to characterize lifetime history of TBI and RHI exposure. The BISQ is a semi-structured questionnaire that queries blows to the head by providing 20 contextual recall cues (e.g., in a vehicular accident, falling from a height). For each reported exposure, subsequent items record the presence and duration of unconsciousness and/or altered mental status, and date of injury. Injury severity classification is made for each exposure per standard criteria.^121^ Head trauma exposure history was verified through review of medical and neuroimaging records whenever available.

### Procedures

Participants attended study visits at ISMMS. Each study visit included a clinical assessment, comprehensive neuropsychological evaluation, blood draw, and MRI scan. This study reports on longitudinal data from the subset of individuals from ISMMS who completed two study visits with blood draw. Demographic, health, and BISQ data were provided by participant self-report at LETBI visit 1. *APOE* genotype was determined from blood samples provided at LETBI visit 1. Blood-based protein data are examined at both visits. All visits for these analyses were conducted between 5/21/2014 to 12/10/2021.

#### Blood analysis

Blood samples are collected in 7mL and 10mL EDTA tubes. Within 30 minutes of blood draw, samples are centrifuged at 1500g for 15 minutes. 1.5mL of plasma is then aliquoted into each cryovial. Aliquoted plasma samples are placed upright in −80°C freezers within 2 hours of collection and stored there until use. Plasma analysis was conducted at the Sahlgrenska Academy at the University of Gothenburg using ultrasensitive Single molecule array (Simoa) technology on the HD-X platform (Quanterix, Billerica, MA, USA). All samples were analyzed using the same batch of reagents by certified laboratory technicians who were blinded to clinical information. The proteins analyzed were: Aβ_42_, Aβ_40,_ GFAP, NfL, and p-tau181 (commercial kits from Quanterix), and BD-tau and p-tau231 using in-house assays as previously described.^129,130^ For some participants, data was not available for one or more proteins(s) as their samples had insufficient volume or were below the limit of quantification (i.e., the lowest concentration of a biomarker that can be reliably measured with acceptable accuracy and precision in each assay).

### Statistical Analysis

Multivariate linear mixed effect models with adjustments for multiple comparisons were performed using the lmerTest Package^131^ in Rstudio^132^ to identify the change in level for 6 unique proteins: Aβ_42_/Aβ_40,_ GFAP, NFL, BD-tau, p-tau231, and p-tau181 between two visits in the same participant. In each model, visit was a fixed effect, participant was a random effect, age and age squared were covariates, and the level of protein in blood was the outcome. Protein data met criteria for Missing Completely at Random (due to insufficient volume/below the limit of quantification), so pairwise deletion was used when data were missing. To correct for multiple comparisons, *p*-values were corrected using false discovery rate, producing *q*-values that were considered significant when *q* < .05.

Spaghetti plots were generated for each protein to visualize heterogeneity and outliers in the data. In addition to observing the spaghetti plots, leverage and sensitivity analyses were completed to explore the extent to which outliers had an undue effect on results. The first sensitivity analysis excluded only the most extreme of outliers to preserve robustness and allow for expected heterogeneity in the data; i.e., outliers defined as 400% of interquartile range beyond the first and third quartile. A more conservative sensitivity analysis was also conducted that removed all outliers defined as 150% of interquartile range beyond the first and third quartile. An additional sensitivity analysis was conducted that removed all participants who had TBIs within a year before their second visit to remove potential bias from pathological response associated with acute injury. A final series of sensitivity analyses with additional covariates were also run to determine whether the relationship over time was still significant over and above the covariate. In addition to using each of these models as sensitivity analyses, chi-square tests with false discovery rate multiple comparison adjustments were used to compare each model against the model with only age and age squared covariates to determine if the model could be improved by addition of the covariate. Demographic covariates were selected based on their known associations with protein levels in TBI and/or non-TBI cohorts (see above): sex, *APOE*4 carrier status, and comorbidities (defined as sum of diabetes, hypertension, heart failure, and stroke; coded as 0 or 1+). Based on emerging evidence about the importance of injury-related factors for blood-based protein levels^133^ we also included the following covariates: age at first blow to the head, years since first blow to the head, and injury severity.

## Results

A total of 63 participants provided blood samples for analysis (Table 1). Participants were 48% female (Age M 52 years; SD 13.41) and were on average 28.04 years (SD 16.3) since their first blow to the head. The most severe injury was classified as severe for 62% of the sample. Between visits there was on average of 3.52 years (SD 1.34; Min 2.04; Max 7.48).

**Table 1.** Demographic characteristics of the sample (n=63)

| Variable | N | % |
| --- | --- | --- |
| Sex |  |  |
| Male | 33 | 52 |
| Female | 30 | 48 |
| *APOE* ε4 carrier status | 16 | 25 |
| Injury severity |  |  |
| Mild | 9 | 14 |
| Complicated Mild | 8 | 13 |
| Moderate | 7 | 11 |
| Severe | 39 | 62 |
|  | Mean | SD |
| Age | 52 | 13.41 |
| Age at first blow to the head | 28.98 | 19.75 |
| Years since first blow to the head | 28.04 | 16.30 |
| Years between visits | 3.52 | 1.34 |

Note. Injury severity refers to the most severe injury recorded; years since first blow to the head is calculated to time of first visit. *APOE* ε4 carrier status is coded as binary (carrier vs non-carrier). SD – standard deviation

### Change over time in proteins

After adjustment for age and age squared and correction for multiple comparisons, two markers exhibited significant change between visits: Aβ_42_/Aβ_40_ and GFAP (Table 2, Figure 1). Aβ_42_/Aβ_40_ ratio tended to decrease by .00383 pg/mL between visits (β = −-0.004, SE = 0.001, t = −3.75, q = .001). GFAP tended to increase by 12.962 pg/mL between visits (β = 12.96, SE = 4.41, t = 2.94, q = .01). There were no significant changes for NfL, BD-tau, p-tau231, or p-tau181.

**Table 2.**
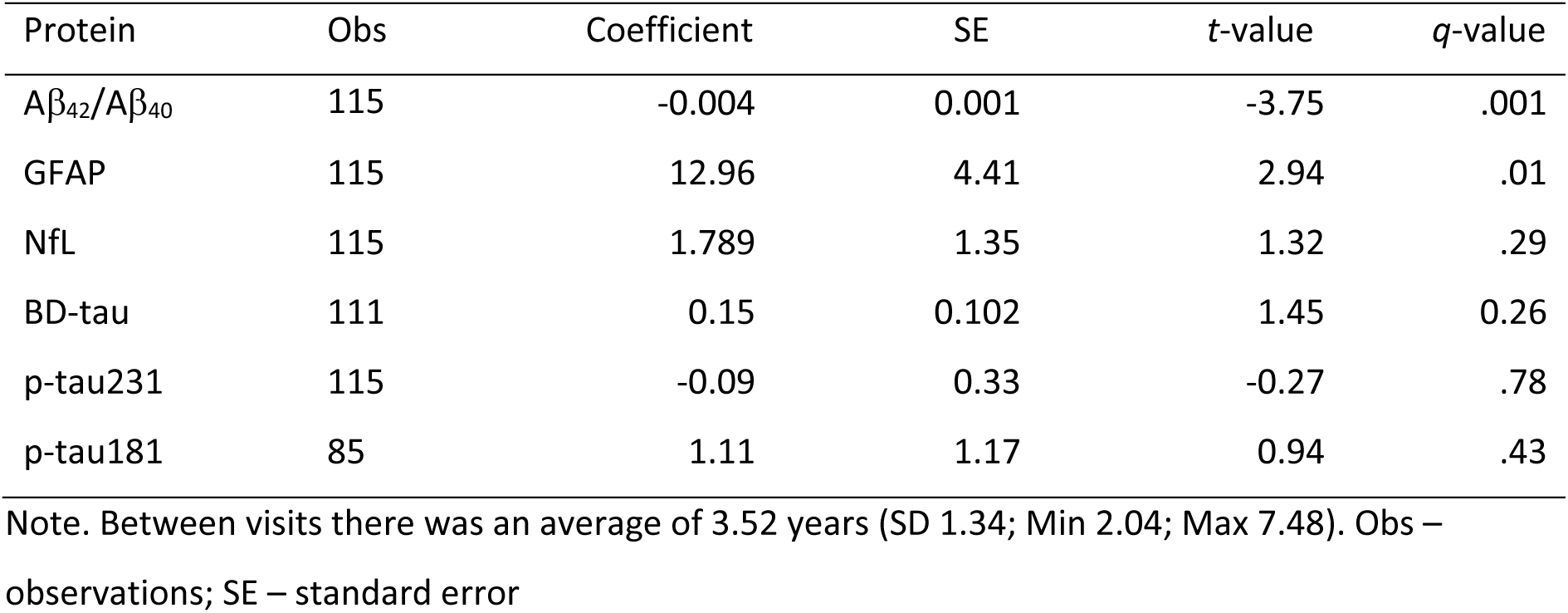
General linear mixed-effect models for proteins across 2 visits.

| Protein | Obs | Coefficient | SE | t-value | q-value |
| --- | --- | --- | --- | --- | --- |
| $A\beta_{42}/A\beta_{40}$ | 115 | -0.004 | 0.001 | -3.75 | .001 |
| GFAP | 115 | 12.96 | 4.41 | 2.94 | .01 |
| NfL | 115 | 1.789 | 1.35 | 1.32 | .29 |
| BD-tau | 111 | 0.15 | 0.102 | 1.45 | 0.26 |
| p-tau231 | 115 | -0.09 | 0.33 | -0.27 | .78 |
| p-tau181 | 85 | 1.11 | 1.17 | 0.94 | .43 |

Note. Between visits there was an average of 3.52 years (SD 1.34; Min 2.04; Max 7.48). Obs – observations; SE – standard error

**Figure 1.**
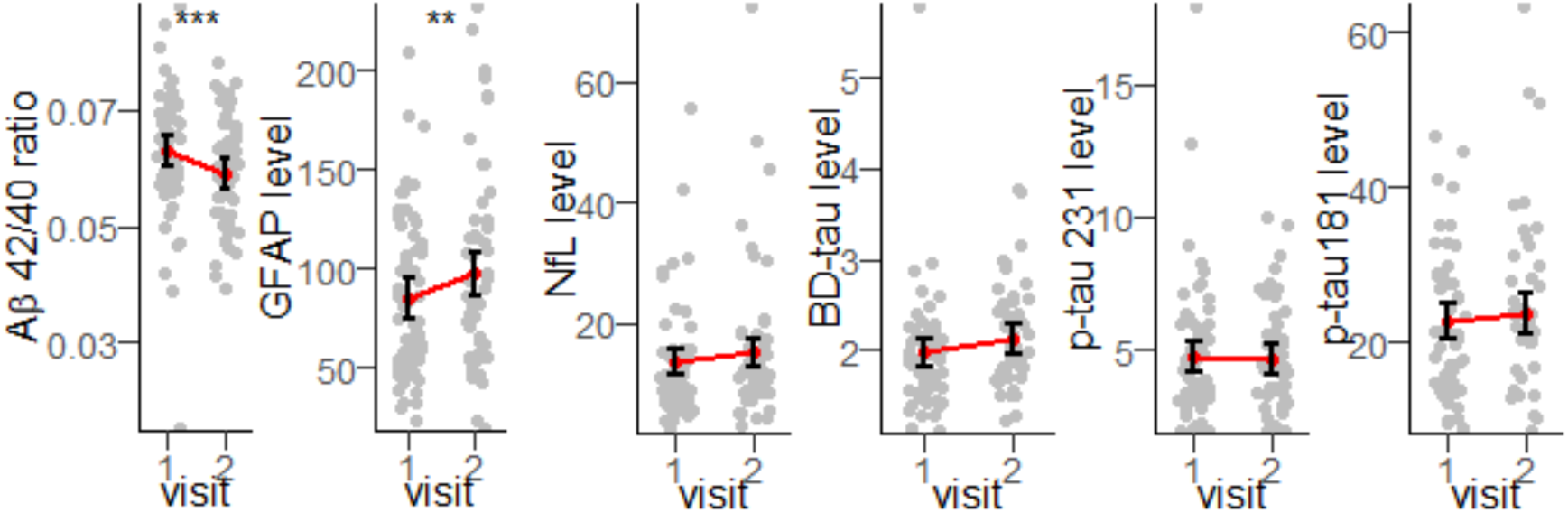
Estimated marginal means, confidence intervals, and change in proteins between visits. Aβ_42_/Aβ_40_, *q* = .001; GFAP *q* = .01; NfL *q* = .29; p-tau231 *q* = .78; p-tau181 *q* = .43

#### Sensitivity Analyses

We conducted a series of sensitivity analyses for the two markers showing significant change over time (Aβ_42_/Aβ_40_ and GFAP). Both markers retained statistically significant change in all sensitivity analyses. Specifically, significance did not change when we excluded (1) extreme outliers only, (2) all outliers, or (3) those who sustained a TBI within 1 year of their second visit (Supplemental Table 1).

We next ran a series of models incorporating additional covariates to determine whether changes in protein levels were accounted for by key covariates: sex, *APOE* ε4 carrier status, medical comorbidities, age at first blow to the head, years since first blow to the head, and injury severity. The models with covariates did not impact the significance for either Aβ_42_/Aβ_40_ and GFAP and did not significantly improve model fit compared to the model with age and age squared only (Supplemental Tables 2-13).

## Discussion

We examined trajectories of blood-based protein biomarker levels over an average of 3.5 years in a cohort of chronic TBI survivors who had sustained their first blow to the head an average of 28 years prior. Consistent with our hypotheses, there were significant reductions in Aβ_42_/Aβ_40_ and significant increases in GFAP over time, implicating a possible role for brain amyloidosis in long-term TBI neuropathology. These findings were robust to key covariates (current age, sex, *APOE* ε4 carrier status, medical comorbidities, age at first blow to the head, years since first blow to the head, injury severity). There were no significant changes in NfL, BD-tau, p-tau231, or p-tau181. To our knowledge, this is the first study to examine change in Aβ_42_/Aβ_40_ and GFAP over time in a large sample with TBI survivors in the very chronic period.

Lower Aβ_42_/Aβ_40_ in blood is an established marker of high Aβ deposition in the brain.^134–140^ This inverse association occurs when Aβ_42_ peptides, which are more prone to aggregation than Aβ_40_,^141^ accumulates into amyloid plaques,^142^ sequestering Aβ_42_ in the brain tissue, leading to decreased levels of soluble Aβ_42_ in blood and lowering the Aβ_42_/Aβ_40_ ratio. Lower Aβ_42_/Aβ_40_ ratio in blood is associated with higher risk of later developing MCI,^143,144^ dementia,^145–147^ AD,^143,144,146^ and greater cognitive decline in healthy control subjects^148^ and community dwelling older adults.^149^ Cross-sectional studies confirm lower Aβ_42_/Aβ_40_ ratio in blood is observed in MCI^136^ and AD^135^ patients compared to controls, and is associated with poorer cognitive performance in these cohorts.^139^

There is preliminary evidence of dysregulated Aβ levels in blood from TBI cohorts. In the acute post-injury period, one study reported increases in Aβ_42_ over the first months post injury in 13 TBI survivors, with elevations remaining for a subset of participants to 3 months post-injury.^150^ A cross-sectional analyses of Aβ in the chronic period showed significant elevations in two neurodegenerative disease-associated Aβ protein variants (A4, C6T) relative to controls in 25 TBI survivors at an average of 30 years post-injury.^57^ In contrast, however, a larger study (n=138) did not identify elevated levels of Aβ_42_ an average of 8.3 years post-injury compared controls using similar covariates to our analyses (covariates: age, body mass index APOE e4 status, gender).^151^ Taken together, it remains unclear whether later dysregulations in Aβ result from a continued acute-phase response or from new trigger(s) in the chronic period after the acute response subsides. Acute elevations in Aβ are well established in TBI and attributed to axonal injury,^36,81,152–154^ with a contribution from increased *APP* gene expression also implicated.^74,153,155^ Maintenance of Aβ elevations in to the chronic post-injury period may be facilitated by persistent axonal injury,^36,45^ self-propagation,^156^ and/or neuroinflammation,^157–159^ which can persist for decades following injury.^160^ Self-propagation may be achieved directly by Aβ stimulating transcription factors that up-regulate Aβ cleavage enzymes,^156^ or indirectly by generating various cellular dysfunctions such as oxidative stress, which, through various feedback loops, have the net effect of up-regulating Aβ.^161^

Consistent with our hypothesis, we found that GFAP levels increased over time in this chronic TBI cohort. GFAP is the signature intermediate filament of astrocytes.^162^ Astrocytes are essential for many functions, including regulating blood flow, supporting synaptic activity, and maintaining extracellular fluid, ion and neurotransmitter homeostasis.^162^ Astrocytes also form an integral physiological and functional part of the blood-brain barrier (BBB).^163^ In response to insults to the central nervous system, astrocytes can become reactive and perform essential beneficial functions, including BBB repair, protection of neurons, and regulation of inflammation.^162^ Elevated GFAP is considered an indicator of astrocytic activation or damage,^162,164^ which are prominent features of TBI.^162,165^ Trajectories of GFAP levels have been previously examined in the post-acute period. A significant increase over time was identified in a small sample (n=12) from approximately 8 months to M 8.3 years post-injury.^166^ However, several other studies have not identified significant changes in GFAP over time (pre-injury to 12 months post-injury;^167^ 3-12 months^98^; 30 days to 5 years^94^). Notably, one of these studies found that GFAP was elevated in the moderate and severe TBI groups compared to controls at each time point (12 months, 2 years, 3 years, 4 years, 5 years),^94^ which may suggest an ongoing but stable elevation. Further cross-sectional studies in the chronic period also support an ongoing elevation, with significantly higher levels in TBI cohorts across severity levels compared to controls (12 months,^92,93^ 6 to 13 years,^168^ M ∼7 years^95^). However, without longitudinal analysis these studies cannot discern whether these cross-sectional snapshots reflect an ongoing but stable elevation or an increase over time.

Elevated GFAP levels have also been identified in other neurodegenerative conditions including frontotemporal lobar degeneration,^169,170^ AD,^171,172^ Dementia with Lewy Bodies,^171^ Parkinson’s disease,^173^ Parkinson’s Disease Dementia,^171^ and cognitively normal older adults at risk of dementia^174^ and AD.^138^ In ADRDs, higher GFAP in the blood has been associated with poorer performance on cognitive tests,^139,170,171^ a steeper rate of cognitive decline,^174,175^ worse functional status,^170^ and higher rates of eventual conversion to dementia.^175,176^ When compared with MRI data, higher GFAP has also been associated with faster decline in hippocampal volume compared to those with lower GFAP.^175^

Ultimately, understanding the complex pathologies of long-term TBI and PTND will require an appreciation of the inter-relationships between pathologies. High GFAP in blood has been robustly related with high Aβ burden.^138–140,176–178^ Up-regulated GFAP may reflect a neuroprotective response whereby reactive astrogliosis occurs as a mechanism of Aβ peptide degradation.^162^ Indeed, astrocytes internalize and degrade Aβ via lysosomal, autophagic, and enzymatic pathways, including the production and expression of Aβ-degrading proteases such as neprilysin.^179^ In contrast, up-regulated GFAP has also been associated with Aβ-burdened reactive astrocytes and astrocyte-derived amyloid plaques that occur when astrocytes internalize but fail to fully degrade Aβ.^180^ Such functionally impaired astrocytes are common in AD.^181^ Complex statistical modelling with multiple data points is required to explore the evolving relationship between Aβ and GFAP in chronic TBI.

Contrary to our hypotheses, changes in NfL, BD-tau, p-tau231, and p-tau181 were not observed. This expands upon prior work in acute TBI samples reporting decreases within the first 12 months for BD-tau^97^ and p-tau231,^97^ suggesting that levels may plateau beyond 12 months post injury. Longitudinal studies report decreases in NfL from 30 days to 5 years^94^ and ∼8 months to M 8.3 years post-injury.^166^ Early decreases captured in these studies may obscure long-term stability, with evidence supporting decline in NfL over the first 12 months post-injury.^97,98^ It is also notable that these studies had smaller sample sizes, with only 29^94^ and 12^166^ TBI survivors included at the final follow-up. Null findings may also reflect the heterogeneity expected in chronic TBI. Firstly, we know that only a sub-set of individuals will experience decline in the post-injury period.^19–21^ For these individuals, whom we would expect proliferating pathology, the constellation of pathologies driving this change may differ inter-individually given preliminary evidence that PTND may be best conceptualized as a poly-pathology.^37,38^ As such, for this study any significant increases in NfL, BD-tau, p-tau231, or p-tau181 may have been obscured by those with (1) other PTND pathologies and/or (2) those without PTND. Indeed, there remain strong theoretical grounds to continue exploring these markers given robust associations with other neurodegenerative diseases and dementia (NfL,^182^ BD-tau,^183^ p-tau231,^184^ and p-tau181^185^).

### Limitations

This study is not without limitations. First, the lack of a matched control group precludes investigation into whether the presence and rate of protein changes differ in relation to those without TBI. Our results were, however, robust to several key covariates (current age, sex, *APOE* ε4 carrier status, medical comorbidities) that may impact proteins levels in the blood. Examining associations with clinical outcomes is a critical next step to identify pathological processes that may contribute to clinical decline observed in some TBI survivors years after injury.^19–21^ It will be of great interest to understand how changes in proteins over time are coupled with clinical manifestations of decline, including the domain(s) affected (e.g. cognitive, neurobehavioral, motor, psychiatric, sleep), the onset, and progression of decline. Although the sample size was small, our study is one of the largest conducted with a cohort of chronic TBI survoivrs.^89^ Larger samples will be required to facilitate analyses of heterogeneity in trajectories that may help to identify sub-groups who have or at risk for PTND.

There are also several considerations inherent to all studies using blood-based biomarkers to make inferences about protein expression in the brain. It is possible the plasma levels of a protein also partly reflects changes in the BBB permeability and/or glymphatic system disruption; which are both thought to occur in TBI and may persist into the chronic period.^186,187^ Further, both GFAP^162^ and Aβ^188^ are generated outside of the central nervous system, with peripheral tissue expression undoubtedly contributing to the overall plasma levels obtained.

## Conclusions

This study provides preliminary evidence for ongoing changes in Aβ_42_/Aβ_40_ and GFAP between study visits conducted an average of 3.5 years apart in individuals living with chronic TBI. Consistent with our hypotheses, we observed significant reductions in Aβ_42_/Aβ_40_ and significant increases in GFAP that were robust to adjustment for key demographic and injury covariates. Hypothesized changes in NfL, BD-tau, p-tau231, or p-tau181 were not observed over this same time interval. Changes in Aβ_42_/Aβ_40_ and GFAP have been previously associated with cognitive decline and risk of dementia, and both are used as biomarkers for brain amyloidosis. Blood-based biomarker analyses have the potential to accelerate our understanding of chronic TBI neuropathology and identify potential pathological process driving PTND.

## Data Availability

Data are available upon reasonable request to the authors.

## Acknowledgements

We acknowledge with thanks to the participants who contribute so generously to the LETBI study. We also acknowledge the LETBI study team and thank them for their continued efforts.

## Appendix A. Supplementary data

## Authorship Contribution Statement

Amelia J Hicks: Conceptualization, Data Curation, Methodology, Project Administration, Supervision, Writing – Original Draft, Writing – Review & Editing.

Jay Plourde: Data Curation, Formal Analysis, Methodology, Software, Validation, Visualization, Writing – Review & Editing

Enna Selmanovic: Investigation, Project Administration, Writing – Review & Editing.

Nicola de Souza: Data Curation, Software, Writing – Review & Editing.

Kaj Blennow: Investigation, Resources, Writing – Review & Editing.

Henrik Zetterberg: Investigation, Resources, Writing – Review & Editing.

Kristen Dams-O’Connor: Conceptualization, Funding Acquisition, Project Administration, Supervision, Writing – Review & Editing.

