## Appendix A. Supplementary data for "Trajectories of blood-based protein biomarkers in chronic traumatic brain injury"

**Supplementary Materials**

**Table 1**

*Model with recent injury (<1 year prior to second visit) removed*

| Protein | Obs | Coefficient | SE | *t*-value | *q*-value |
| --- | --- | --- | --- | --- | --- |
| Aβ_42_/Aβ_40_ | 112 | -0.00271 | 0.000738 | -3.67 | 0.002 |
| GFAP | 112 | 9.882968 | 3.122926 | 3.16 | 0.006 |

Note. 3 people were removed; Obs – observations; SE – standard error

**Table 2**

*Nested model with sex covariate*

| Protein | Obs | Coefficient | SE | *t*-value | *q*-value |
| --- | --- | --- | --- | --- | --- |
| Aβ_42_/Aβ_40_ | 115 | -0.00388 | 0.001024 | -3.79125 | <.001 |
| GFAP | 115 | 13.16882 | 4.413536 | 2.983734 | .004 |

Obs – observations; SE – standard error

**Table 3**

*Model comparison of sex covariate*

| Protein | chisquare | q |
| --- | --- | --- |
| Aβ_42_/Aβ_40_ | 3.226546 | .14 |
| GFAP | 2.144307 | .38 |

**Table 4**

*Nested model with age at first injury covariate*

| Protein | Obs | Coefficient | SE | *t*-value | *q*-value |
| --- | --- | --- | --- | --- | --- |
| Aβ_42_/Aβ_40_ | 114 | -0.00383 | 0.001023 | -3.74007 | <.001 |
| GFAP | 114 | 13.34562 | 4.380133 | 3.046852 | 0.003 |

Obs – observations; SE – standard error

**Table 5**

*Model comparison of age at first injury covariate*

| Protein | chisquare | q |
| --- | --- | --- |
| Aβ_42_/Aβ_40_ | 0.126537 | .87 |
| GFAP | 1.294883 | .38 |

**Table 6**

*Nested model with years since first injury covariate*

| Protein | Obs | Coefficient | SE | *t*-value | *q*-value |
| --- | --- | --- | --- | --- | --- |
| Aβ_42_/Aβ_40_ | 114 | -0.00383 | 0.001023 | -3.73993 | <.001 |
| GFAP | 114 | 13.37525 | 4.379414 | 3.054119 | .003 |

Obs – observations; SE – standard error

**Table 7**

*Model comparison of time since first injury covariate*

| Protein | chisquare | q |
| --- | --- | --- |
| Aβ_42_/Aβ_40_ | 0.016933 | .90 |
| GFAP | 1.478662 | .38 |

**Table 8**

*Nested model with APOE e4 genotype covariate*

| Protein | Obs | Coefficient | SE | *t*-value | *q*-value |
| --- | --- | --- | --- | --- | --- |
| Aβ_42_/Aβ_40_ | 115 | -0.00383 | 0.001019 | -3.76239 | <.001 |
| GFAP | 115 | 13.01394 | 4.400314 | 2.957502 | 0.004 |

Note. APOE coded as carrier vs non-carrier. Obs – observations; SE – standard error

**Table 9**

*Model comparison of APOE ε4 genotype covariate*

| Protein | chisquare | q |
| --- | --- | --- |
| Aβ_42_/Aβ_40_ | 3.226546 | 0.144907 |
| GFAP | 0.126537 | 0.866461 |

Note. *APOE ε4* coded as carrier vs non-carrier

**Table 10**

*Nested model with injury severity covariate*

| Protein | Obs | Coefficient | SE | *t*-value | *q*-value |
| --- | --- | --- | --- | --- | --- |
| Aβ_42_/Aβ_40_ | 115 | -0.00393 | 0.001024 | -3.83823 | <.001 |
| GFAP | 115 | 12.75791 | 4.41591 | 2.889079 | .005 |

Note. Injury severity coded as mild, mild complicated, moderate, and severe

Obs – observations; SE – standard error

**Table 11**

*Model comparison of injury severity covariate*

| Protein | chisquare | q |
| --- | --- | --- |
| Aβ_42_/Aβ_40_ | 4.628305 | .09 |
| GFAP | 0.803517 | .44 |

Note. Injury severity coded as mild, mild complicated, moderate, and severe.

**Table 12**

*Nested model with medical comorbidities*

| Protein | Obs | Coefficient | SE | *t*-value | *q*-value |
| --- | --- | --- | --- | --- | --- |
| Aβ_42_/Aβ_40_ | 115 | -0.00385 | 0.001022 | -3.76891 | <.001 |
| GFAP | 115 | 12.95468 | 4.406403 | 2.939968 | .004 |

Note. medical comorbidities were diabetes, hypertension, heart failure, and stroke and coded as binary 0 or 1+; Obs – observations; SE – standard error

**Table 13**

*Model comparison of comorbidities (0 or 1+)*

| Protein | chisquare | q |
| --- | --- | --- |
| Aβ_42_/Aβ_40_ | 1.952261 | .24 |
| GFAP | 0.039211 | .84 |

Note. Medical comorbidities were diabetes, hypertension, heart failure, and stroke and coded as a binary 0 or 1+
